# The Effect of Electrode-Tissue Distance on Unipolar and Bipolar Voltage Electrograms for large-Tip, Ring-, and Mini-Electrodes

**DOI:** 10.1101/2024.01.24.24301751

**Authors:** Vincent Schlageter, Adrian Luca, Patrick Badertscher, Philipp Krisai, Thomas Küffer, David Spreen, Josip Katic, Stefan Osswald, Beat Schaer, Christian Sticherling, Michael Kühne, Sven Knecht

**Affiliations:** Department of Cardiology, University Hospital Basel, University of Basel, Basel, Switzerland; Cardiovascular Research Institute Basel, University Hospital Basel, University of Basel, Basel, Switzerland; Department of Cardiology, Lausanne University Hospital and University of Lausanne, Lausanne, Switzerland; Department of Cardiology, Inselspital, Bern University Hospital, University of Bern, Bern, Switzerland

**Author notes:** Corresponding author: Sven Knecht, PD DSc University Hospital Basel Petersgraben 4 4031 Basel, Switzerland.

## Abstract

**Background:** The characteristics of intracardiac unipolar and bipolar voltage electrograms (EGM) acquired by electrophysiological catheters depend on the electrode design and configuration.The aim of the study was to assess the impact of electrode design and distance from the myocardial electric source on the unipolar and bipolar intracardiac electrograms recorded with a multi-electrode ablation catheter do deduce a cut-off for far-field versus near-field discrimination.

**Methods:** We retrospectively analyzed left atrial electroanatomical maps of 25 patients performed using an ablation catheter with a 4.5 mm tip-, mini- and 2 mm ring electrodes. The unipolar and bipolar EGMs were characterized based on peak-to-peak amplitude, signal duration (width), maximal slope, and relative power of the high frequency spectrum (HF_rel). Distances of the electrode from the tissue were calculated from the electroanatomic reconstruction.

**Results:** We analyzed EGMs of 5183 catheter positions. The unipolar EGM of ring electrodes showed an increased amplitude (140%), slope (150%) and HF_rel (16% vs 11%) compared to the tip- and mini-electrodes. In contrast, for bipolar EGM, the tip-ring pair showed the largest amplitude, width, and slope. The median amplitude, slope, and HF_rel for the ring electrodes followed a power-law decay function with distance. A cut-off of 4 mm was determined for far-field versus near-field discrimination.

**Conclusions:** We showed a higher unipolar amplitude for small ring-electrodes compared to larger tip electrodes. Furthermore, a rapid decay of the amplitude, slope, and HF_rel with distance could be observed. The decay functions are suggestive for a near-field cut-off distance below 4mm from the tissue.

## Introduction

The characteristics of unipolar and bipolar voltage electrograms (EGM) acquired by electrophysiological catheters depend on the electrical source as well as on the sensing electrode configuration. The resulting EGM is used to identify the timing of the electrical activation and to characterize the myocardial substrate to perform catheter ablation. Despite following this basic principle for decades, the exact relationship between electrical source and resulting unipolar voltage (UV) and bipolar voltage (BV) EGM is still not completely understood. The UV EGM reflects the far-field and near-field information of the cellular depolarization wavefront traveling along the electrode. The BV EGM, however, calculated as the difference between two unipolar electrodes, eliminates a large fraction of the far-field information and mainly reflects the local near-field information.^1,2^ However, the BV EGM strongly depends on the electrode pair orientation relative to the electrical activation wavefront. This limitation was recently addressed to some extent by the introduction of multipolar diagnostic catheters and dedicated software allowing to calculate a orientation-independent BV EGM.^3,4^ The impact of the distance between the electrical source and the electrodes on the EGM characteristics in general, however, is still not known. This is of high clinical relevance to reliably characterize the underlying substrate and to be able to successfully plan and perform safe and effective catheter ablation procedures. For the latter, knowledge if the electrical source of a sensed electrogram can be reached by the energy of the ablation catheter is of utmost importance. Finally, despite being used in daily clinical practice, the cut-off to differentiate between far-field and near-field EGM has never been quantitatively specified.

Hence, the aim of this study was to assess the impact of the electrode type and distance from the electric source on the bipolar and unipolar intracardiac EGM recorded with a multi-electrode ablation catheter and to deduce a potential cut-off for far-field versus near-field discrimination.

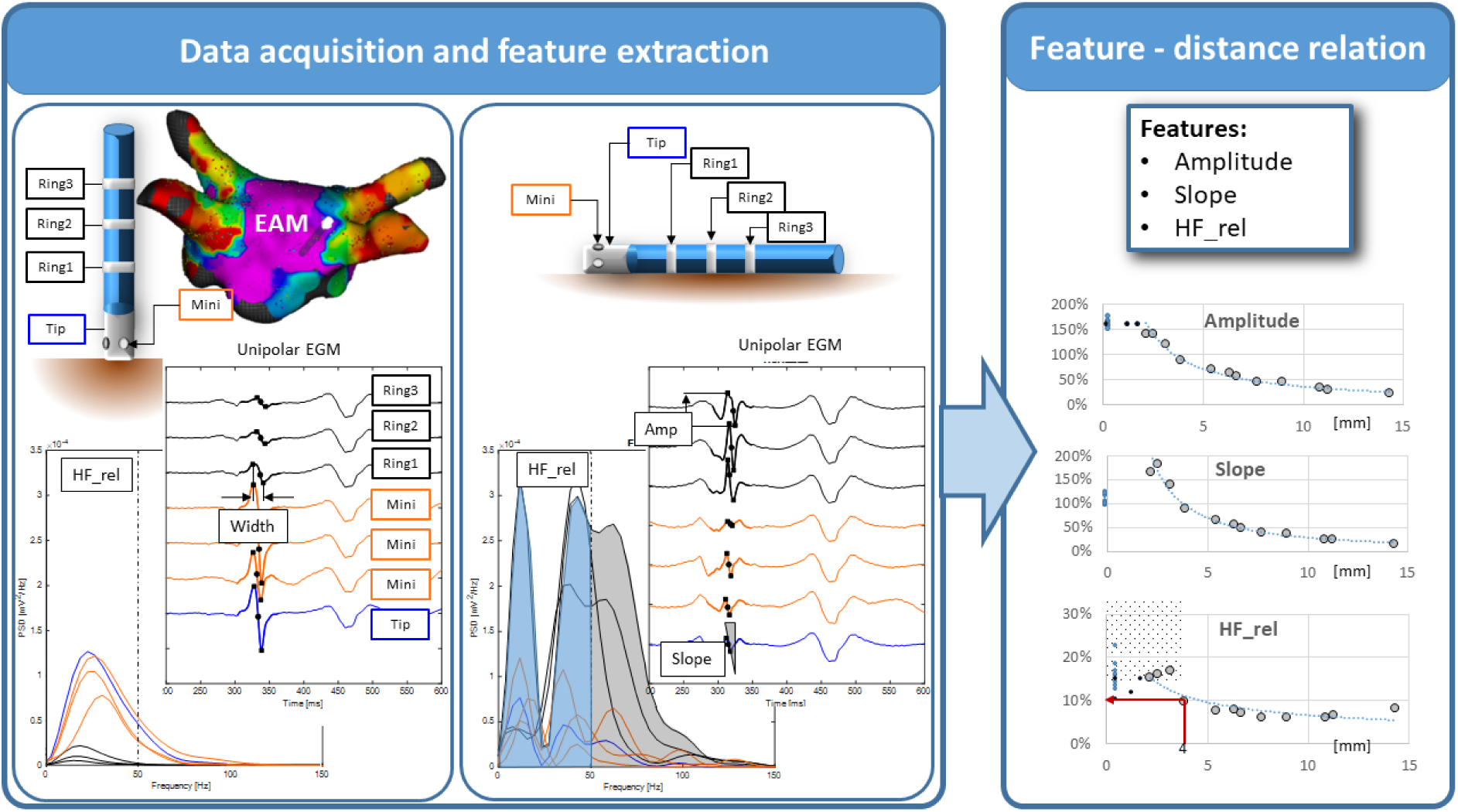

Central Illustration: From the detailed electroanatomic map (EAM), the unipolar and bipolar electrograms were extracted (only unipolar shown here). Based on the catheter orientation, the distance of the electrodes to the tissue was calculated. The amplitude, the slope, the width and the relative power of the high-frequency spectrum above 50 Hz (HF_rel, grey area in middle block of the horizontal catheter orientation) were calculated for all electrodes (tip-, ring-, and mini-electrodes). Representative feature-distance relationship for the unipolar voltage are shown in the right column.

## Methods

We analyzed the left atrial (LA) 3D electroanantomical maps (EAMs) from the prospective CHAZE study (ClinicalTrials.gov Identifier: NCT04095559). In this study, 25 patients with atrial fibrillation underwent detailed mapping of the LA in sinus rhythms prior to repeat catheter ablation. The study was approved by the local ethics committee (Ethics Committee Northwest and Central Switzerland) and was conducted in accordance with the Declaration of Helsinki.

### Electrogram acquisition

The LA EAM was acquired in sinus rhythm using the IntellaNav Mifi OI catheter as previously described using the Rhythmia system (Boston Scientific, USA).^5^ In brief, the LA anatomy was mapped using the IntellaNav Mifi OI catheter (7.5 Fr, 4.5 mm tip electrode size, mini electrode size 0.5 mm^2^ with 2.4 mm interelectrode spacing and 1.3mm from tip; 2mm ring electrode with 2.5mm interelectrode spacing). Electrode surfaces of the tip-, ring- and mini-electrode measure 30 mm^2^, 10 mm^2^ and 0.5 mm^2^, respectively. The bipolar voltage (BV) and unipolar voltage (UV) were filtered at 10-250 Hz with a sampling rate of 1 kHz.

### Electrogram Analysis

The exported study data included the location and BV and UV EGMs of all three electrode types (tip-, ring-, and mini-electrode) and the anatomical shell The data was processed and analyzed in Matlab (MATLAB. (2021). Natick, Massachusetts, USA) as follows: The catheter orientation relative to the reconstructed 3D electroanatomical shell was calculated and categorized into four groups: parallel (0° to 14°), medium low (15° to 34°), medium high (34° to 59°), and perpendicular (60°to 90°). The stratification was employed to cover a wide range of electrode distances from the tissue (Figure 1). In perpendicular orientation the theoretical source-to-electrode distance at the center-of-mass is half the tip size (2.3 mm) for the tip electrode, 1.3 mm for the MIFI electrodes and 8 mm, 12.5 mm, 17 mm for the first, second, and third ring electrode, respectively. In parallel orientation, the theoretical distance for the center-of-mass of the electrode is half the catheter diameter (1.25 mm). The effective distance was calculated based on the position of the electrode and the projection to the surface shell of the EAM (Figure 1). The impact of the electrode dimension on the EGM characteristics was deduced from the parallel orientation group, whereas the medium and perpendicular orientation groups were used to deduce the distance dependency of the electrodes.

**Figure 1:**
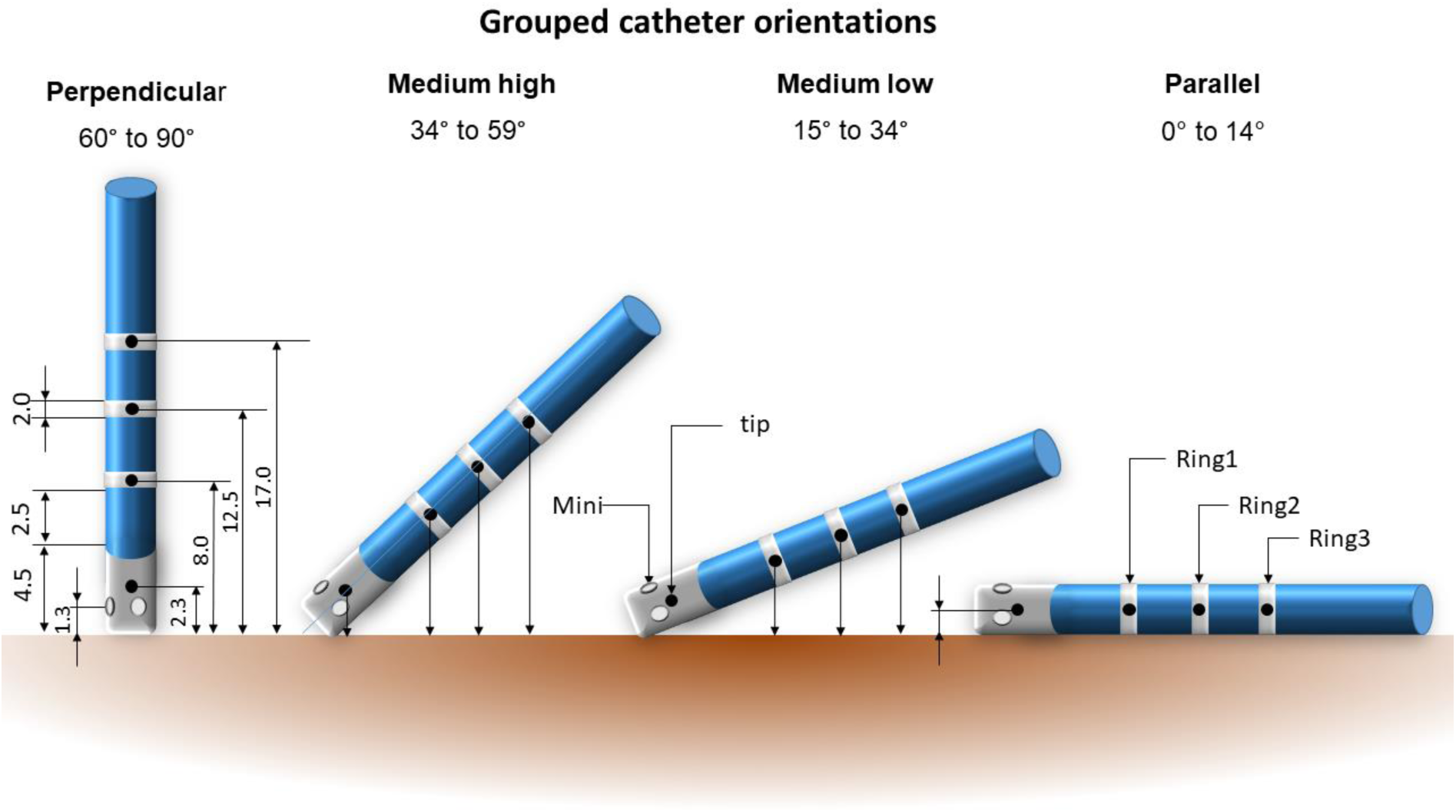
Catheter orientation, dimension, and distance to tissue The perpendicular and tilted catheter orientations were used to investigate the impact of electrode type and distance on the EGM. In contrast, the parallel catheter orientation was used to investigate the impact of electrode configuration only due to the comparable electrode-tissue distance. The black dots represent the center-of-mass of the electrodes for which the distances were reported based on the projection to the tissue.

The EGMs were quantified based on the peak-to-peak amplitude (amp), the duration between min and max value (width), the maximal slope dV/dt between the minimal and maximal values, and the relative power of the high-frequency spectrum above 50 Hz (HF_rel) (Central Illustration). The latter was calculated using fast Fourier transform (FFT) as recently published.^6^ Briefly, the power of the high-frequency spectrum was calculated for a frequency cut-off of 50 Hz (50-300 Hz) and normalized by the power spectrum of the entire spectrum. Except for the HF_rel, for which absolute values are reported, all features are normalized using the tip-electrode for unipolar or the tip-to-ring pairs for bipolar measures as a reference.

### Statistical Analysis

Continuous variables are expressed as median with IQR. To investigate the relationship between the above-described features and the distance from the electrical source, we plotted all features over the distance between the electrode center-of-mass and the myocardial surface and performed a regression analysis. The best model was selected based on the highest R-squared statistic.

## Results

We analyzed EGMs of 5183 catheter locations based on the four features amplitude, width, slope, and HF_rel from 25 patients (72% male, age 68±15 years, LVEF 59±8%, LA size 41±8 mm, LAVI 38±9 ml/m^2^). Thereof, 1345 EGMs were from parallel, 1826 from medium low, 1588 from medium high and 421 EGMs from perpendicular catheter orientation. The median value of the BV tip-ring amplitude was 0.79 mV (IQR: 0.41 mV, 1.55 mV).

### Impact of electrode size on the EGM characteristics

Based on the analysis of the parallel catheter orientation with all electrodes being within similar proximity with the tissue, the smaller ring electrodes showed an increased unipolar amplitude (140%), increased slope (150%) and higher HF_rel (Table 1) compared to the tip electrode. Furthermore, the signal width was smaller for the ring compared to the tip electrode. For the mini-electrodes, amplitude, slope and width are only slightly increased compared to the tip electrode. For the bipolar EGMs, however, the tip-ring pair showed the largest amplitude, width, and slope. The HF_rel component was larger for all electrode pairs compared to the corresponding unipolar EGM, but lowest for the tip-ring1 electrode pair.

**Table 1:**
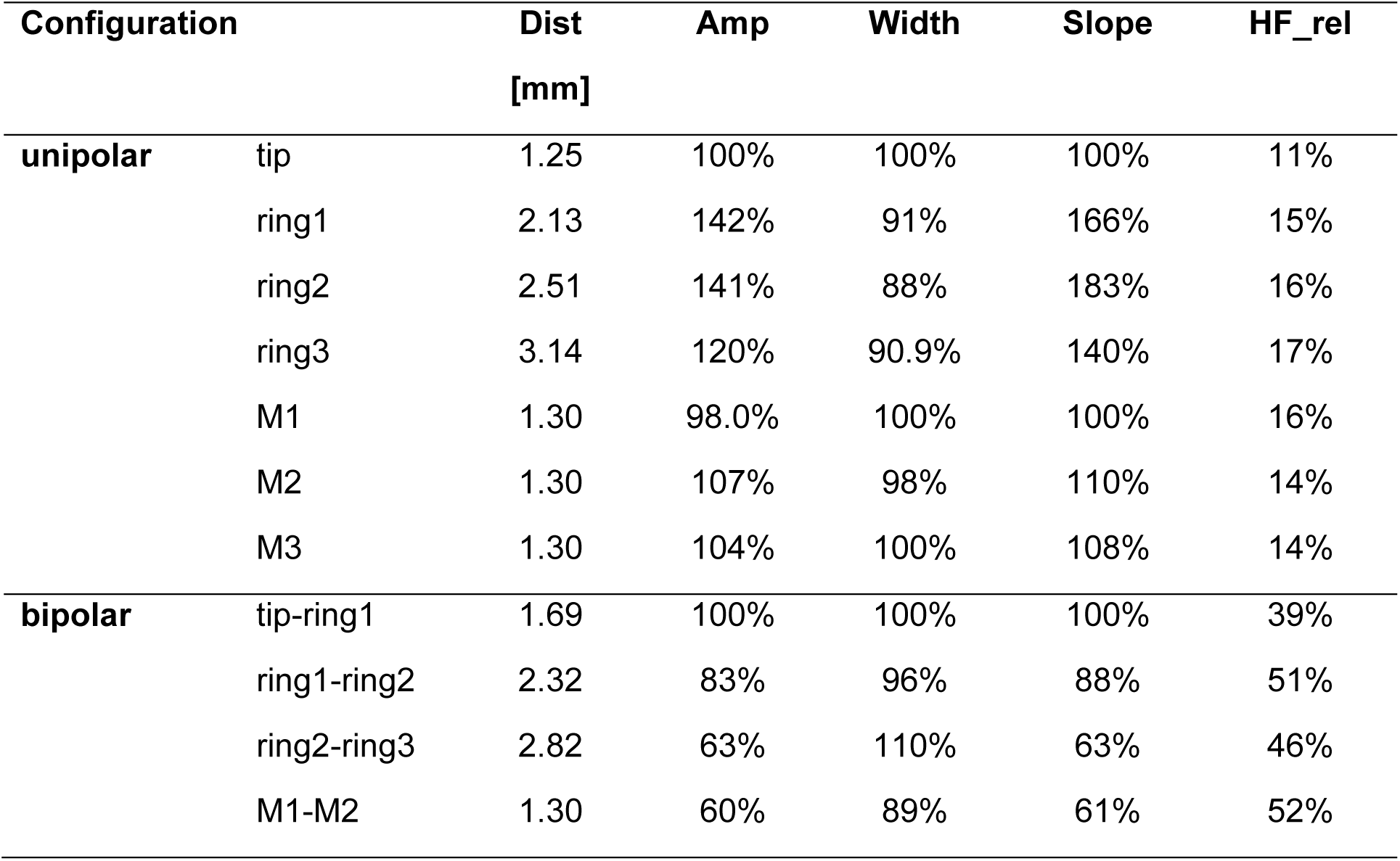
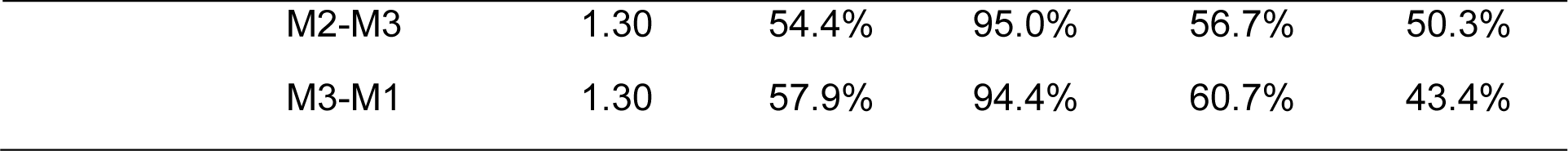
Parameters for the uni- and bipolar EGMs for the three electrode types for the parallel orientation. Values of amp, width and slope are normalized using the tip-electrode for unipolar and the tip-to-ring pairs for bipolar signals. Amp - voltage amplitude, Dist – distance from tissue, M – mini-electrode.

| Configuration |  | Dist<br>[mm] | Amp | Width | Slope | HF_rel |
| --- | --- | --- | --- | --- | --- | --- |
| **unipolar** | tip | 1.25 | 100% | 100% | 100% | 11% |
|  | ring1 | 2.13 | 142% | 91% | 166% | 15% |
|  | ring2 | 2.51 | 141% | 88% | 183% | 16% |
|  | ring3 | 3.14 | 120% | 90.9% | 140% | 17% |
|  | M1 | 1.30 | 98.0% | 100% | 100% | 16% |
|  | M2 | 1.30 | 107% | 98% | 110% | 14% |
|  | M3 | 1.30 | 104% | 100% | 108% | 14% |
| **bipolar** | tip-ring1 | 1.69 | 100% | 100% | 100% | 39% |
|  | ring1-ring2 | 2.32 | 83% | 96% | 88% | 51% |
|  | ring2-ring3 | 2.82 | 63% | 110% | 63% | 46% |
|  | M1-M2 | 1.30 | 60% | 89% | 61% | 52% |

Table 1: Parameters for the uni- and bipolar EGMs for the three electrode types for the parallel orientation.
|  |  |  |  |  |  |
| --- | --- | --- | --- | --- | --- |
| M2-M3 | 1.30 | 54.4% | 95.0% | 56.7% | 50.3% |
| M3-M1 | 1.30 | 57.9% | 94.4% | 60.7% | 43.4% |

In a perpendicular catheter orientation with the mini-electrodes being all in comparable distance from the tissue within around 1.3 mm, their variation of all unipolar features was relatively small (absolute difference of 2%). In parallel orientation, however, with one electrode being potentially in full contact with the tissue whereas the other electrodes are at 2.5 mm distance based on the catheter diameter, a larger variation was observed especially in the amplitude and slope of 8-10%. Of note, the ring electrodes showed a strong dependency on the catheter orientation in all features, whereas the values of the tip electrodes with a stable contact to the tissue remained comparable (supplemental figure).

### Relationship between ECG features and distance

The electrode specific relationship between the distance and the EGM features are shown in Figure 2 for unipolar and Figure 3 for bipolar EGMs. The three features amp, slope, and HF_rel followed a power-law decay function (R^2^=0.97, R^2^=0.95 and R^2^=0.79, respectively), whereas the EGM width increases linearly with distance from the tissue (R^2^=0.96). Since the catheter tip was required to be in contact with the tissue during mapping, the information on distance-dependency of the tip and mini-electrode is limited. The decline for the three features is less pronounced for the bipolar amplitude compared to the unipolar amplitude, representing the local characteristics of the bipolar EGM.

**Figure 2:**
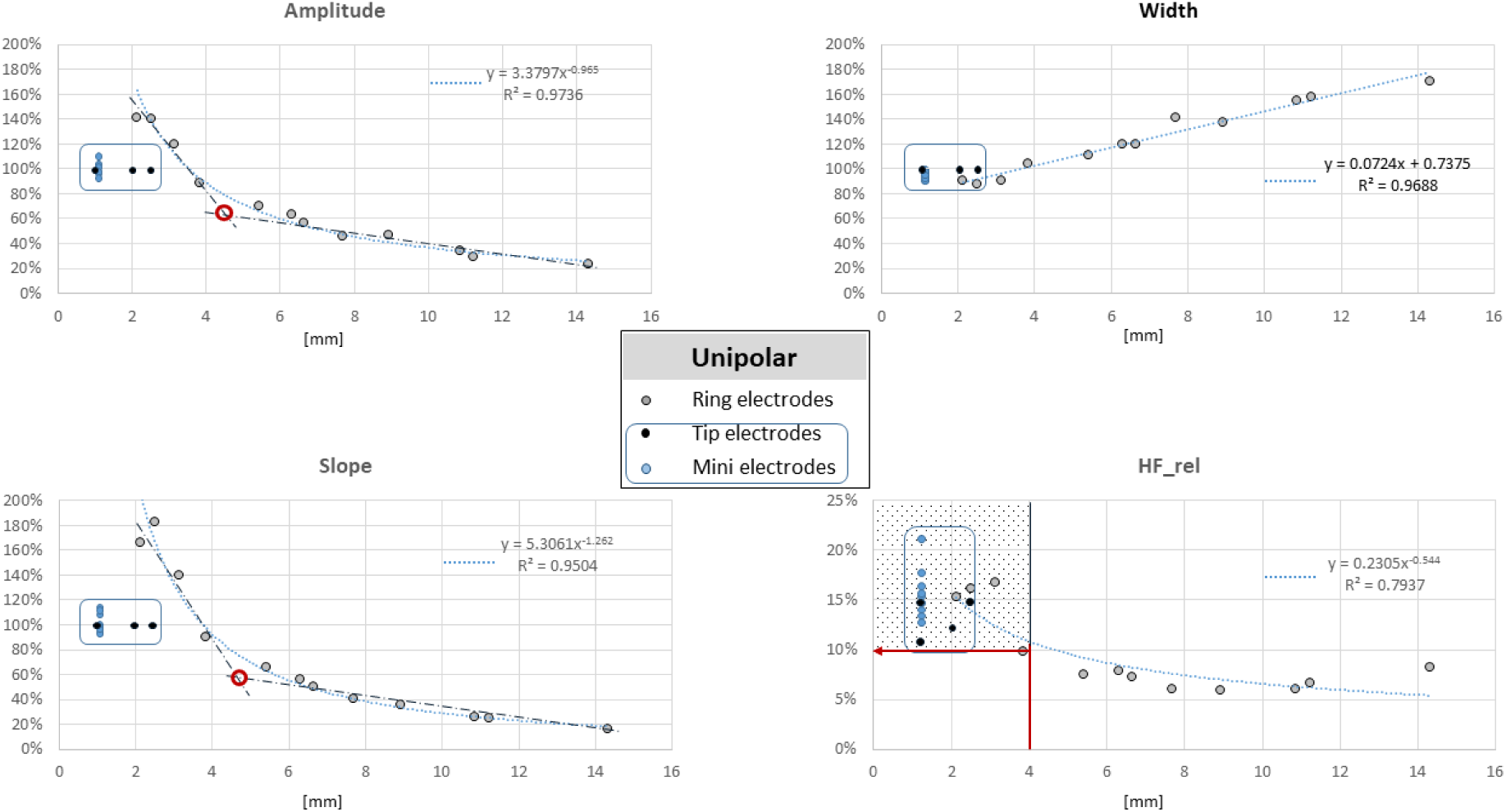
Feature-distance plot of the unipolar EGM for the amplitude, width, slope, and relative power of the high-frequency spectrum (HF_rel). The blue dotted line reflects the best fit. The blue line encloses the features obtained from the catheter tip (tip-, and mini-electrodes).

**Figure 3:**
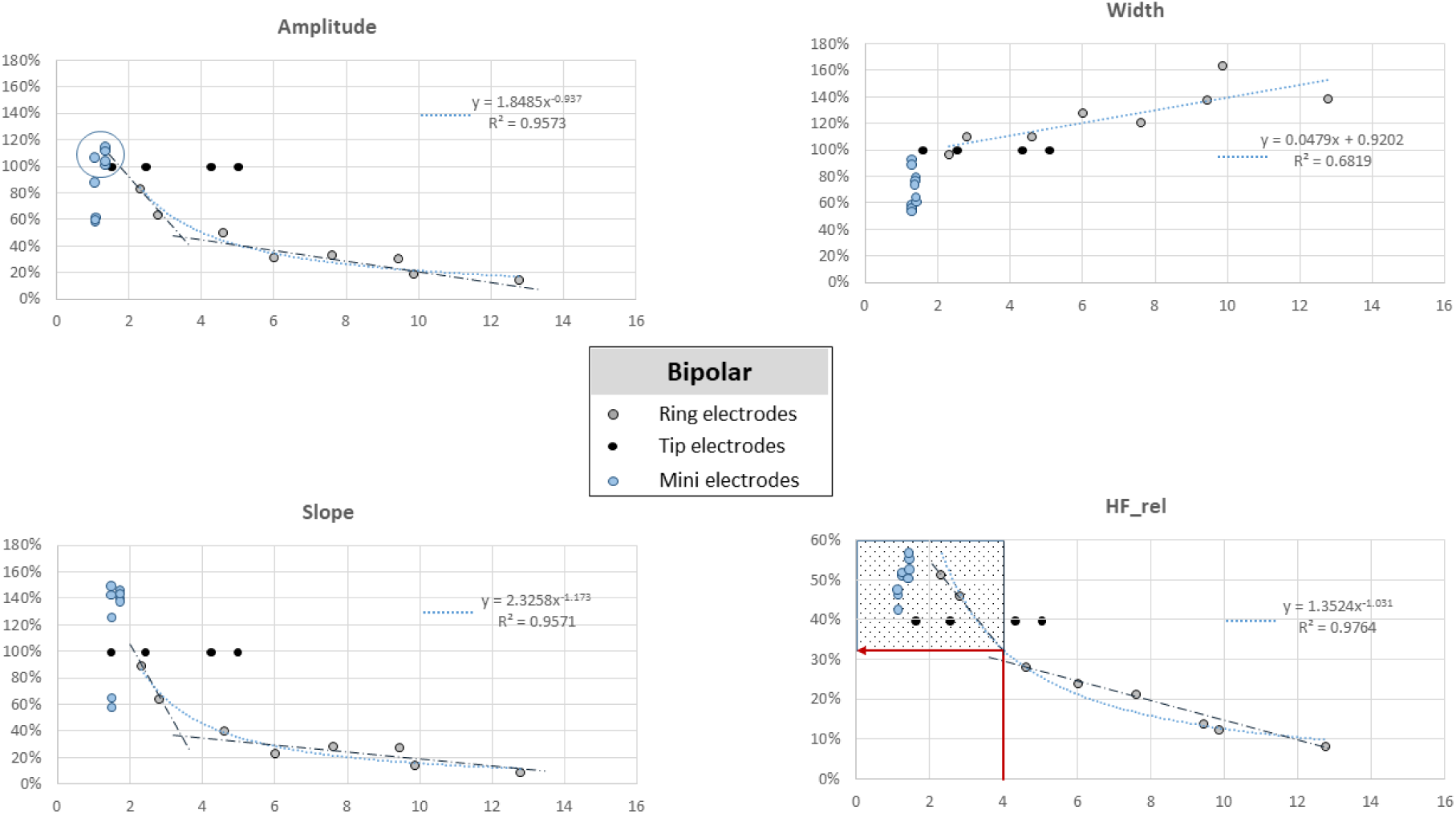
Feature-distance plot of the bipolar EGM for the amplitude, width, slope, and relative power of the high-frequency spectrum (HF_rel). The blue dotted line reflects the best fit.

The gradients of the fitted curves for the two features amp and slope gradually decrease between 4 mm and 6 mm distance, as reflected by the angulation of the two linear approximations (dash-dotted line, Figure 2 & 3) for close and distant points at their intersection (red circles, Figures 2 & 3). Accordingly, this distance up to 4 mm was defined as near-field, which was confirmed for the bipolar EGMs. At this cut-off, a HF_rel value of >10% for unipolar (Figure 1, red arrow) and >30% for bipolar EGMs (Figure 2, red arrow) was observed.

## Discussion

Unipolar and bipolar EGMs are often characterized qualitatively based on their visual appearance as near-field or distant far-field EGM source. Except for numerical simulations, however, the relationship of EGM characteristics and distance from the tissue to deduce a distance-EGM relationship has not been characterized quantitatively. In our detailed analysis of EAMs of the LA, we could observe the following: 1) The ring electrodes with a smaller surface area than the tip showed a larger amplitude, slope, and high frequency component HF_rel and a narrower signal width. For bipolar EGM, however, the ring electrode pairs showed a lower amplitude, slope, and HF_rel compared to the distal tip-ring electrode. 2) The mini-electrodes showed unipolar features comparable to the tip electrode. However, based on the higher variation of the unipolar signals in parallel compared to perpendicular orientation, a strong relationship with distance might be characteristic for this electrode type. 3) The amplitude, slope, and HF_rel for the ring electrodes showed all a strong power-law decay function with distance, whereas the EGM width increases linearly with distance from the tissue. This could be observed for the unipolar and for the bipolar EGMs. 4) A steep linear decline for a distance up to 4 mm is suggestive for a strong near-field relationship, whereas the flat decay above 6 mm is suggestive of mainly far-field impact. Based on this value of 4 mm, a near-field cut-off for the unipolar and bipolar electrogram with a HF_rel of 10% and 30%, respectively, appears reasonable.

### Impact of electrode size

It is commonly considered that the smaller the electrode is, the larger the amplitude of the unipolar EGM should be.^7^ We could confirm this assumption in our study for the ring electrodes but not for the mini electrodes, which showed similar UV amplitude to the tip electrode. This might be explained at least in parts by the spatial distribution of the mini electrode on the tip of the catheter. Whereas one electrode is in direct tissue contact in the parallel catheter orientation, the other mini electrode might be more than 2 mm away from the tissue. Due to the missing information of the exact mini-electrode position in the 3D EAM data, however, we could report the UV features for all electrodes for the same location of 1.3 mm only. Further *in vivo* comparisons to investigate the impact of electrode size on the EGM are rare. The comparison of another micro-electrode ablation catheter (Qdot, Biosense) with a multipolar diagnostic catheter (Pentaray, Biosense) in porcine infarction models revealed a moderate linear relationship between the BV EGMs of the two catheters. ^8^ Whereas in healthy ventricular tissue, the smaller micro-electrodes showed larger bipolar electrograms compared to the ring-electrodes from the Pentaray or the tip electrode of the ablation catheter, this difference diminished in diseased, fibrotic tissue. In our preceding study comparing the bipolar EGM in repeat procedure after index PVI, we even identified lower BV amplitude for the catheters with smaller electrode surface, such as the Orion or the circular Lasso catheter compared to the larger tip electrode.^5^ The reason might be the interplay between interelectrode spacing in BV EGMs, spatial fibrosis distribution and amplitude of the electrical source, as well as wavefront propagation direction.

Using UV EGMs from a multipolar catheter (Octaray, Biosense Webster), the area of interest with scar tissue could be better delineated than using the bipolar electrogram.^9^ However, a two-layer viable myocardium (with midmyocardial scar) could be identified more reliable using the BV EGM. The used EGM classification based on local, high-frequency, multicomponent signals was performed qualitatively based on the physicians judgement.^10,11^ Recently, however, frequency-based analysis to characterize and classify the LV substrate were published.^12,13^ Instead of manual annotation of local abnormal ventricular activity, late potentials (LP), or deceleration zone (DZ), this area of fractionation could be identified with a sensitivity of 91% and specificity of 85% by applying a 220 Hz frequency cut-off value in areas of low bipolar voltage based on the HD grid bipolar voltage map.^12^ The smaller ring-electrodes with a length of 1 mm and diameter of 0.8 mm might explain the higher frequency cutoff compared to our applied 50Hz value. Furthermore, our aim was to discriminate near-field from far-field signal and not to identify areas with abnormal local EGM in low voltage areas based on a machine learning algorithm. Prediction of midmyocardial fibrosis from late-iodine computed tomography scans based of detailed frequency-based EGM analysis was performed in 13 patients with non-ischemic cardiomyopathy using a 3.5mm tip ablation catheter.^13^ They reported that frequency-feature based classifiers better predict the presence of midmyocardial fibrosis. In conclusion, both studies confirm the potential of frequency feature extraction from intracardiac EGM to gain a deeper insight into and understanding of the origin of the sensed unipolar and bipolar EGM.

### Impact of electrode distance from tissue

Over a distance up to 14 mm from the tissue, we observed a power-law decay for the voltage, slope, and the fraction of high frequency components HF_rel. The decay function property is in line with the results from a surgical study in patients with paroxysmal and persistent AF.^14^ Based on the experimental data of 10 sec unipolar EGMs from a high-density electrode array and a computer simulation, they observed a decay to 50% of the source unipolar EGM within 1.5 mm to 2.1 mm function. This is in the same range for the ring electrode in our study between 2 mm and 4 mm distance (140% to 70% for amplitude and 180% to 90% for the slope).

Of note, within the close distance of 4 mm of the ring electrodes from the tissue, we observed a linear decline of the amplitude and slope of the unipolar and bipolar EGMs, whereas the signal width remains constant. Furthermore, within this boundary, the fraction of high-frequency components above 50% (HF_rel) in the overall EGM is higher than at more distant positions. This observation reinforces the usage of these features for far-field discrimination.

### Implications for clinical far-field and near-field discrimination

Despite being mentioned and used in numerous studies on signal analysis and interpretation, the term near-field and far-field is not yet quantified in the field of EP. Currently, the EGMs are mainly classified based on the qualitative signal characteristics, such as fractionation, sharpness, or high frequency EGMs.^10,11^ Whereas fractionation can be quantified and reproduced, a signal “sharpness” or presence of “high-frequency” components are still undefined.

To classify the EGM into near-field and far-field based on a specified boundary depends on one hand on the technical background (e.g. source and electrode characteristics) but as well on the ability to reach and eliminate the target EGM by ablation. With recent ablation technologies, lesion depth around 4 to 5 mm can be created.^15,16^ Based on this procedural criterion, the deduced cut-off at 4 mm might be comprehensible. This distance might be represented for all electrode types in the healthy tissue above 0.5 mV by a relative HF component with a 50 Hz above 10% and 30% based on the unipolar and bipolar EGM, respectively. To verify these values, however, further prospective studies investigating as well the application on smaller tip electrode sizes of commonly 3.5 mm are recommended.

## Limitations

First, the acquired 3D-EAM might not reflect the actual location of the myocardium, undermining calculated catheter-to-tissue distances. This effect is particularly pronounced at short distances, where a minor deviation can lead to a significant percentage error. Second, the observed relationship and deduced cut-off are only valid for atrial tissue. Especially for the ventricle with thicker tissue and consequently larger voltage and differences in frequency composition, difference in values must be expected. In parallel orientation, the catheter might cover not purely the same tissue with the same electrical source. In conclusion, difference between tip and ring might arise as well from differences of the source and not purely of the electrodes. We analyzed the electrodes being surrounded by the bloodpool with a different conductivity than the tissue, which might have an impact on the sensed EGM. Finally, we could not investigate the influence of catheter distance for the tip electrode since all mapped points were with the tip in contact with the tissue. Whether the cutoffs deduced from the ring electrodes are transferable to the tip electrode with different size needs further investigation.

## Conclusion

A strong relationship of the unipolar and bipolar EGMs with electrode size and distance could be observed with a higher amplitude for small ring-electrodes compared to larger tip electrodes. Furthermore, a decay of the amplitude, slope, and HF_rel with distance could be observed. This decay functions are suggestive for a near-field cut-off distance below 4mm from the tissue, which is reflected by a high HF_rel of 10% and 30% for the unipolar and bipolar EGM, respectively. Further studies are warranted to confirm this observation.

## Data Availability

Data will be shared based on reasonable request

## Acknowledgements

None

## Sources of founding

The study was supported by an investigator sponsored grant (Boston Scientific, ISRRM10392).

---

**Supplemental Figure 1:**
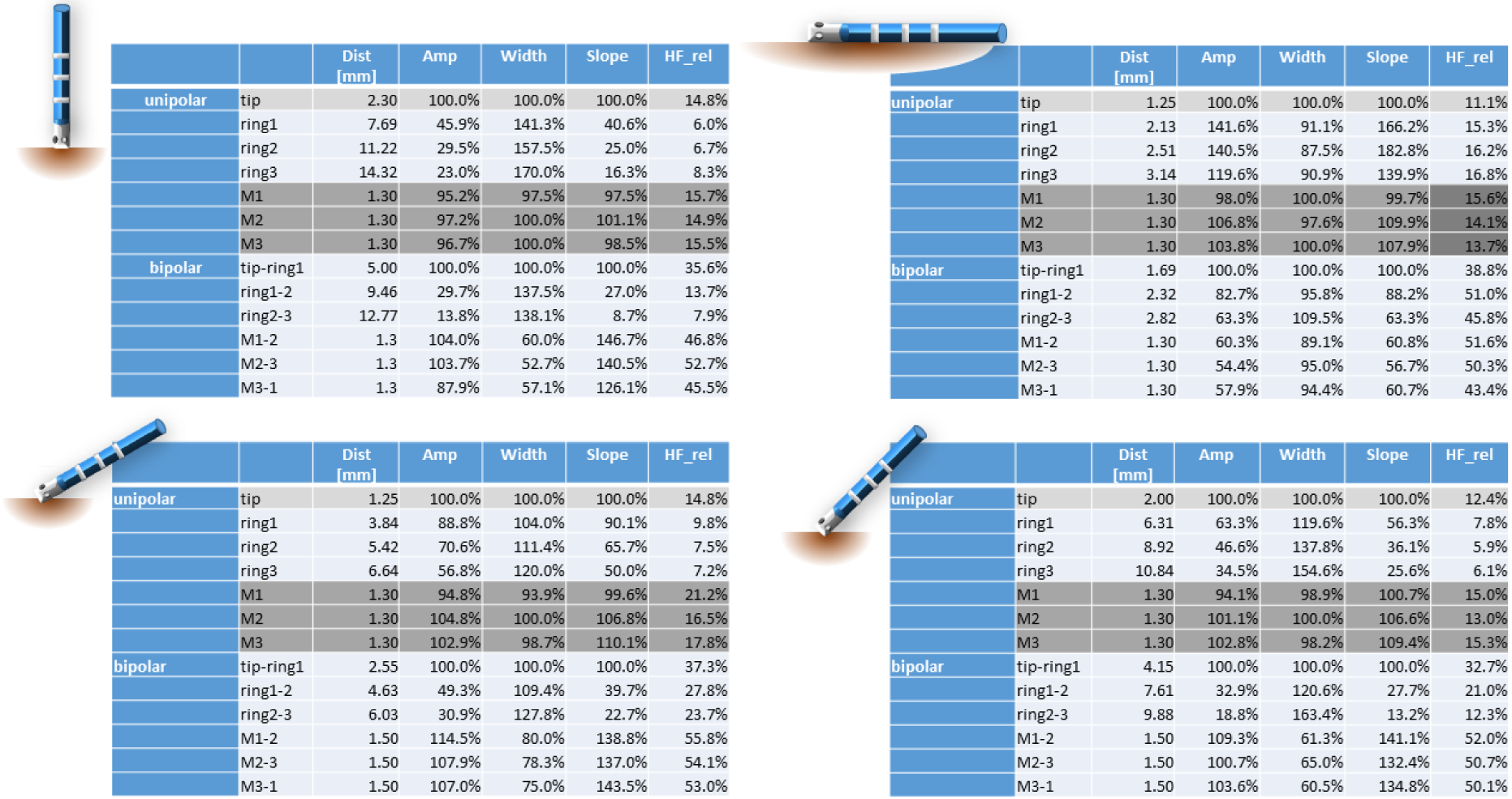

